# Untangling factors associated with country-specific COVID-19 incidence, mortality and case fatality rates during the first quarter of 2020

**DOI:** 10.1101/2020.04.22.20075580

**Authors:** Lev Shagam

## Abstract

At early stages of the COVID-19 pandemic which we are experiencing, the publicly reported incidence, mortality and case fatality rates (CFR) vary significantly between countries. Here we aim to untangle factors that are associated with the differences during the first quarter of the year 2020. Number of performed COVID-19 tests has a strong correlation with country-specific incidence (p < 2 × 10^−16^) and mortality rate (p = 5.1 × 10^−8^). Using multivariate linear regression we show that incidence and mortality rates correlate significantly with GDP per capita (p = 2.6 × 10^−15^ and 7.0 × 10^−4^, respectively), country-specific duration of the outbreak (2.6 × 10^−4^ and 0.0019), fraction of citizens over 65 years old (p = 0.0049 and 3.8 × 10^−4^) and level of press freedom (p = 0.021 and 0.019) which cumulatively explain 80% of variability of incidence and more than 60% of variability of mortality of the disease during the period analyzed. Country hemisphere demonstrated significant correlation only with mortality (p = 0.17 and 0.036) whereas population density (p = 0.94 and p = 0.75) and latitude (p = 0.61 and 0.059) did not reach significance in our model. Case fatality rate is shown to rise as the outbreak progresses (p=0.028). We rank countries by COVID-19 mortality corrected for incidence and the factors that were shown to affect it, and by CFR corrected for outbreak duration, yielding very similar results. Among the countries where the outbreak started after the 15th of February and with at least 1000 registered patients during the period analyzed, the lowest corrected CFR are seen in Israel, South Africa and Chile. The ranking results should be considered with caution as they do not consider all confounding factors or data reporting biases.

## Introduction

The coronavirus disease 2019 (COVID-19) that emerged in China in late 2019 and is caused by the SARS-CoV-2 virus from the *Coronaviridae* family (Andersen et al. 2020) has led to almost 47 thousands registered deaths during the first quarter of 2020 only and is having a huge impact on the world economy.

Examination of factors influencing COVID-19 epidemiological parameters and comparison of the latter across countries has crucial importance during and following the pandemic. Here we focus on COVID-19 incidence, mortality and case fatality rate (CFR) and examine their correlations with selected variables characterizing demographic, healthcare, financial, geographical and political aspects of the countries.

Specifically, mortality and case fatality rate for COVID-19 have been previously shown to be associated with age, comorbidities and sex (Onder, Rezza, and Brusaferro 2020). Some studies suggest that country-specific CFR and incidence can depend on latitude (Braiman 2020) and temperature (Triplett 2020), respectively. Apart from testing the abovementioned correlations in our study, here we also examine the following assumptions: (i) number of laboratory tests for the infection could correlate with incidence, mortality or CFR, (ii) COVID-19 incidence at the beginning of pandemic could be associated with the countries’ economic situation (for instance, being impacted by travel activity of the citizens) as well as with population density (potentially facilitating spread of the virus), (iii) finally, incidence, mortality and thus CFR could be misreported in the countries with limited press freedom.

All the three abovementioned epidemiological parameters vary substantially across countries during the period of interest. Comparison of the countries’ healthcare systems in regard to CFR could shed light on best practices of pandemy management (Khafaie and Rahim 2020). It is being extensively discussed though that reported CFR could be biased due to a number of factors. Countries might use different death reporting policies. Hospitalization and laboratory testing capacities can also vary both across and within countries (Onder, Rezza, and Brusaferro 2020) which would create inaccuracies within the data that are very difficult to account for. Also, the CFR which can only be computed using censored data during the course of outbreak significantly depends on the way it is calculated. Assessment based on total number of cases underestimates whereas calculation based on closed cases significantly overestimates the number (Spychalski, Blazynska-Spychalska, and Kobiela 2020). According to WHO, the time between symptom onset and death ranges from about 2 to 8 weeks (WHO, 2020), however dividing the number of deaths by total number of cases 2 weeks before (Baud et al. 2020) turned out to be biased too, especially during the first days of the pandemic (Spychalski, Blazynska-Spychalska, and Kobiela 2020). Here we show that country-specific CFR depends on the number of days that passed from the outbreak start. For comparison of the CFR across countries we correct the fatality rate for duration of the epidemy.

We do not consider potentially different malignancy that can be achieved by mutations of the virus as well as health profile and inherited susceptibility differences across the populations which also could affect all the epidemiological parameters being compared.

## Materials and methods

Data on number of confirmed cases of coronavirus as well as number of deaths per country was obtained as of April 1, 2020 (Dong, Du, and Gardner 2020). Data on press freedom ranking across the globe for the year 2019 was obtained from (Reporters without borders, 2019). Population, area, GDP per capita and percentage of population over the age of 65 data was taken from (World Bank Open Data, 2020). Information on number of performed COVID-19 tests per country was obtained from (Our world in data, 2020) (when the number was not known for April 1st, it was extrapolated using the information for adjacent dates).

We collected and used the following country-specific covariates for analysis: daily records of number of COVID-19 cases as well as deaths, country centroid latitude, population size, area, press freedom ranking measure, number of tests performed and fraction of people over 65 years old.

Calculations were performed in R version 3.6.0 using packages stats, tibble and data.table, visualization was done using package ggplot2. COVID-19 incidence was calculated as a ratio of total number of detected cases by a certain date to total population of the country. Mortality was calculated as a ratio of total number of COVID-19-related deaths by a certain date to total population of the country. For some calculations and plots (as indicated below), incidence and mortality rate were subjected to base-ten logarithmic transformation. Case fatality rates were calculated as country-specific ratios of total deaths to total number of diagnosed cases for all days of monitoring till the date of interest (typically 01 April, 2020) and then subjected to base-ten logarithmic transformation. P-value significance cut-off across all tests was set at 0.05. Performing univariate regression was preferred over multivariate whenever the number of countries with data available was less than 40-50.

We analyzed data for a total of 201 countries or territories (overseas territories of France, United Kingdom and Netherlands were considered as separate in our analysis) with at least one patient reported. In order to increase accuracy of the case fatality analysis, we considered only countries/territories with at least 1000 registered COVID-19 patients which was a total of 49 for this specific part of the analysis. Countries known to perform extensive healthy population testing for COVID-19 during the period of interest (Iceland) were excluded from the analysis.

## Results

### Countries cluster into two groups based on start date of the infection process

For each country, we calculated time from the moment when the number of registered cases of COVID-19 was at least 10 till the 1st of April, 2020 (also referred to as duration of the outbreak/epidemic below). As it is shown on fig. 1, the distribution of the epidemic duration times divides countries into two groups: with duration of ≤40 and of ≥53 days. For reason of uniformity, the group of 12 countries with outbreak start date before 15th of February (see fig. 1a) was excluded from further statistical analysis.

**Figure 1.**
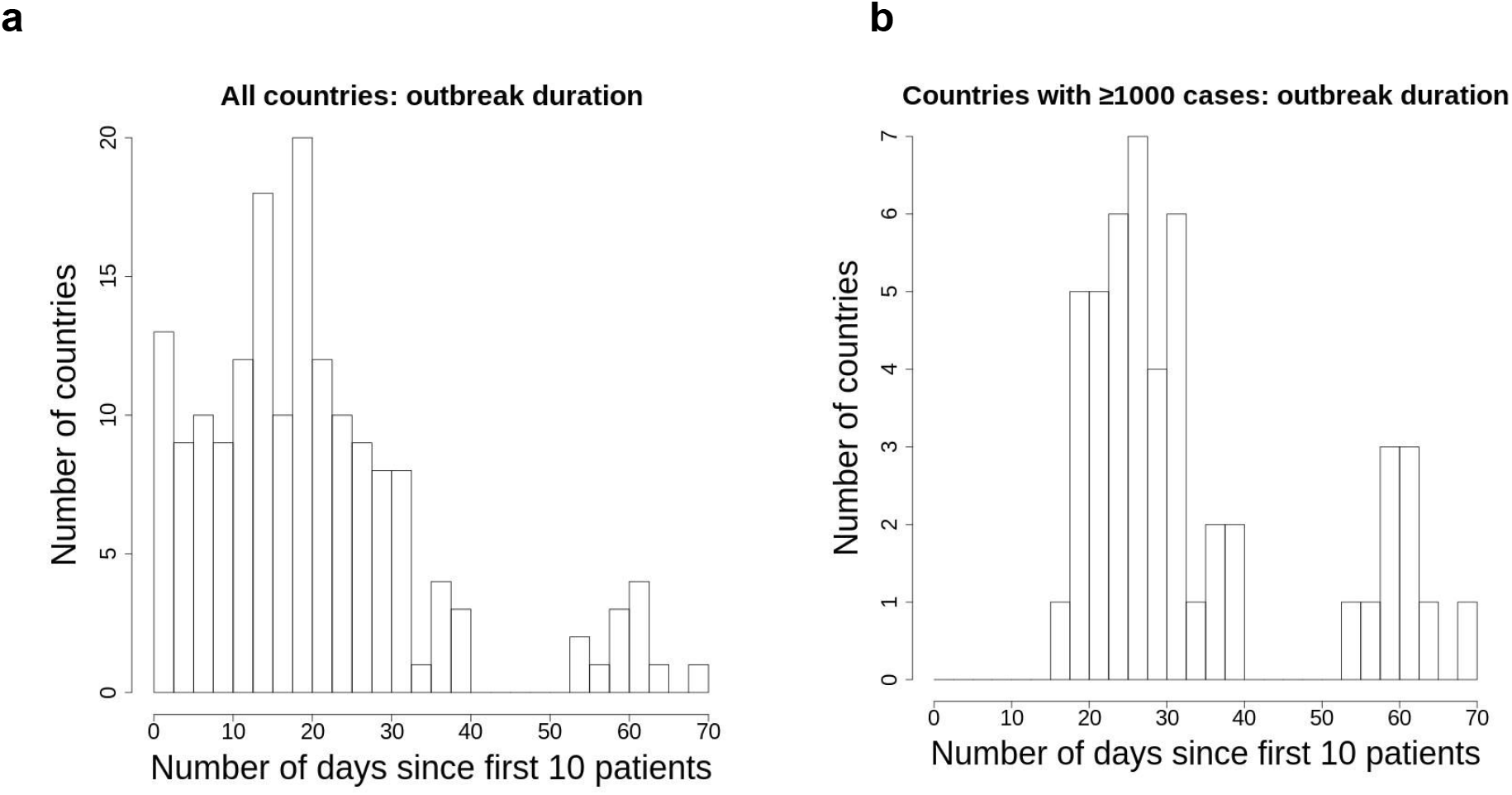
Number of days from first 10 registered cases of COVID-19 till April 1st, 2020 across countries; **a**, all countries with at least 10 reported cases, **b**, all countries with at least 1000 cases as of April 1st, 2020. Based on the observed bimodal distributions of epidemic durations, countries can be divided into 2 groups with sizes of 156 and 12 (a); 39 and 10 countries each (b).

### Number of tests for COVID-19 correlates with GDP per capita

Number of performed tests is an important factor that could both determine the reported number of cases or mortality and be influenced by the two indicators via actions of medical authorities and testing facilities. However, testing activity report is yet provided by limited number of countries. In a multivariate linear regression model, we checked its correlation (log-transformed) with log-transformed GDP per capita, latitude, hemisphere (Northern or Southern), press freedom ranking measure, population density, log-transformed fraction of people over the age of 65 and outbreak duration (N = 51). The number of tests turned out to strongly correlate with GDP per capita (p = 7.7 × 10^−8^) whereas other variables were not influencing it (total R^2^_adj_ = 0.796).

### Variables explaining COVID-19 incidence and mortality across countries

We examined the dependence of COVID-19 incidence and mortality for the period of January-March, 2020 on some country-specific variables that could potentially influence them. Considering the reasons of avoiding multicollinearity as well as significant sample size reduction mentioned above, this was done using two linear regressions:

i. the model of log-transformed dependent variable explained by country log-transformed GDP per capita, latitude, hemisphere (Northern or Southern), press freedom ranking measure, population density, log-transformed fraction of people over the age of 65 and outbreak duration (R^2^_adj_ = 0.796 for incidence and R^2^_adj_ = 0.625 for mortality, N = 124 and 99; countries with no registered COVID-19 cases or deaths were excluded from the corresponding analyses). Results are summarized in table 1 (columns 2 and 3) and fig. 2 and include among others a correlation of all the three epidemiological parameters examined with duration of the outbreak;

**Table 1.** COVID-19 epidemiological parameters’ correlation with country-specific explanatory variables for the countries with outbreak start after the 15th of February, 2020: regression coefficients (for significant correlations only) and p-values.

| Country-specific variable | Incidence ($\log_{10}$)<br>(N = 124,<br>multivariate<br>regression) | Mortality ($\log_{10}$)<br>(N = 99,<br>multivariate<br>regression) | CFR ($\log_{10}$)<br>(N = 38, univariate<br>regressions) |
| --- | --- | --- | --- |
| Outbreak duration, days | $\beta = 0.021$,<br>$p = 2.6 \times 10^{-4}$ | $\beta = 0.030$,<br>$p = 0.0019$ | $\beta = 0.022$<br>$p = 0.028$ |
| GDP per capita, USD ($\log_{10}$) | $\beta = 9.1$,<br>$p = 2.6 \times 10^{-15}$ | $\beta = 0.53$,<br>$p = 7.0 \times 10^{-4}$ | $p = 0.96$ |
| Latitude (abs. value) | $p = 0.61$ | $p = 0.059$ | $p = 0.50$ |
| Hemisphere (N/S) | $\beta = 0.075$<br>$p = 0.17$ | $\beta = 1.77$<br>$p = 0.036$ | $p = 0.63$ |
| Press freedom ranking measure | $\beta = -7.8$,<br>$p = 0.021$ | $\beta = -0.012$,<br>$p = 0.019$ | $p = 0.71$ |
| Population density | $p = 0.94$ | $p = 0.75$ | $p = 0.10$ |
| Fraction of population over 65 years old ($\log_{10}$) | $\beta = 0.53$,<br>$p = 0.0049$ | $\beta = 1.0$,<br>$p = 3.8 \times 10^{-4}$ | $p = 0.33$ |

**Fig. 2.**
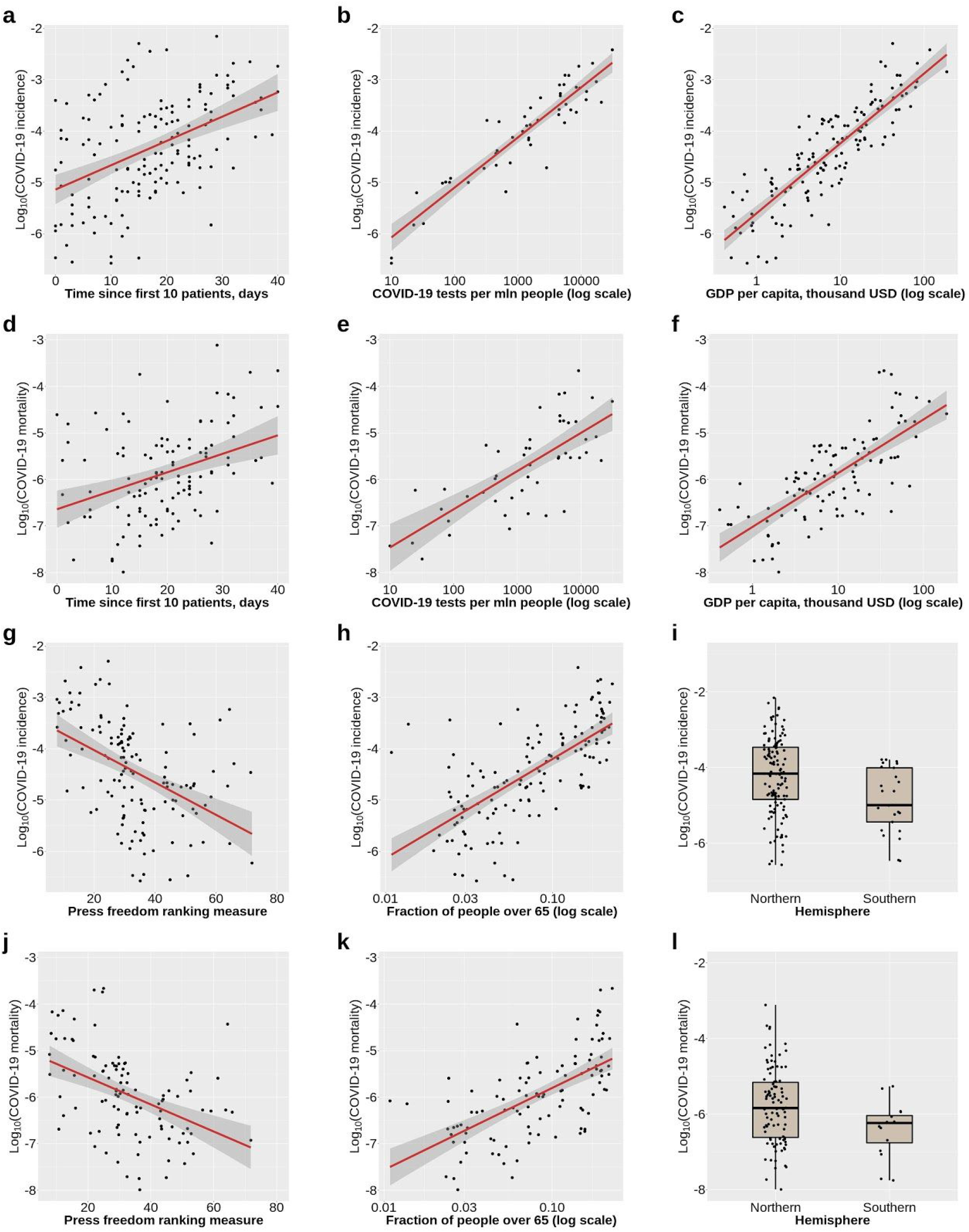
Country-specific COVID-19 incidence (rows 1 and 3) and mortality rate (rows 2 and 4) depending on explanatory variables: outbreak duration (**a**,**d**), number of tests performed (**b**,**e**), GDP per capita (**c**,**f**), press freedom ranking measure (countries with limited press freedom have higher rank; **g**,**j**), fraction of senior citizens (**h**,**k**) and position of the country’s centroid (**i**,**l**). Each dot corresponds to a country (only countries with outbreak start after 15th of February plotted). Linear approximation in the corresponding univariate model (red) as well as 95% confidence interval (grey) are shown.
ii. the model of log-transformed dependent variable corrected for outbreak duration explained by log-transformed number of tests (R^2^_adj_ = 0.741 for incidence, N = 51, and R^2^_adj_ = 0.460 for mortality, N = 49, results are summarized in table 2, columns 2 and 3).

**Table 2.**
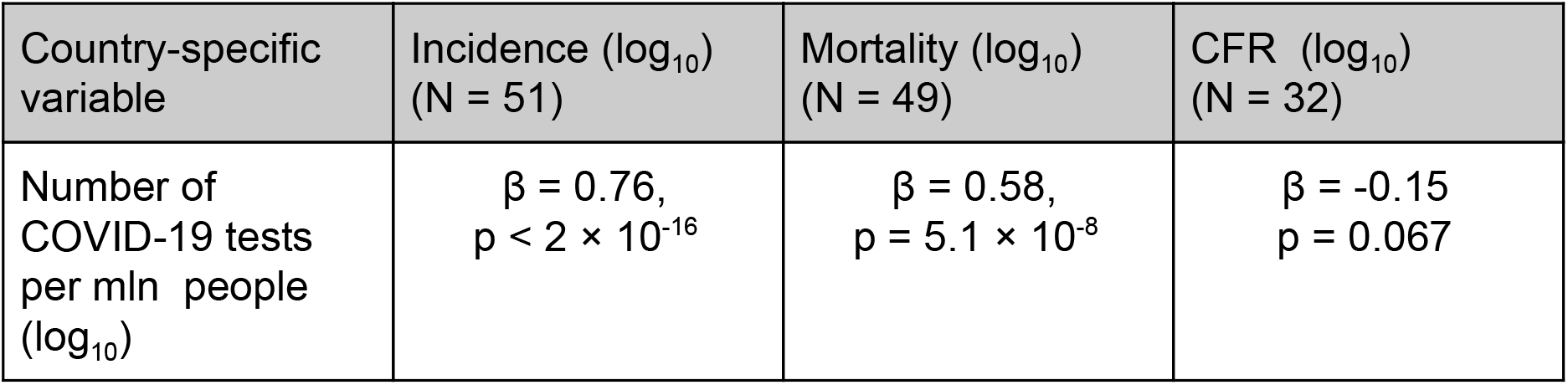
COVID-19 epidemiological parameters’ correlation with number of performed tests for the countries with outbreak start after the 15th of February, 2020: regression coefficients and p-values. Incidence, mortality and CFR were corrected for outbreak duration.

| Country-specific variable | Incidence ($\log_{10}$)<br>(N = 51) | Mortality ($\log_{10}$)<br>(N = 49) | CFR ($\log_{10}$)<br>(N = 32) |
| --- | --- | --- | --- |
| Number of COVID-19 tests per mln people ($\log_{10}$) | $\beta = 0.76$,<br>$p < 2 \times 10^{-16}$ | $\beta = 0.58$,<br>$p = 5.1 \times 10^{-8}$ | $\beta = -0.15$<br>$p = 0.067$ |

GPD per capita, fraction of senior citizens, outbreak duration, press freedom rank are correlated with both incidence and mortality. Hemisphere of the country centroid is associated with mortality, however its association with incidence is not significant. The association of population density as well as of country centroid latitude with neither incidence nor mortality has reached significance in our analysis when other explanatory variables were considered. Number of tests is strongly correlated with both incidence and mortality.

Given the correlation of GDP per capita with number of performed tests as well as a positive feedback loop of increasing testing capacity as number of patients increases, the true causality between these three variables is difficult to dissect. However, we further aimed to shed light on it by understanding if the influence of GDP per capita on the reported incidence and mortality is mediated solely by the number of tests performed or vice versa the observed influence of number of tests is a byproduct of correlation with GDP. We explore this by comparing the additional variance explained by each of the components.

The model of incidence explained by GDP per capita and country-specific epidemy duration explains 82.5% of variation (R^2^_adj_). Addition of number of tests explains 88.5% of variation (p = 4.9 × 10^−3^; 0.071; 6.0 × 10^−6^ for GDP, epidemy duration and number of tests correspondingly), whereas the model of incidence by number of tests and epidemy duration alone explains 86.7% of variation (N = 51). At the same time, when mortality is modeled by the three variables, influence of number of tests becomes not significant (p = 0.54) compared to GDP per capita (p = 0.0092) and time since first ten patients were reported (p = 0.0093). Together these three variables explain 66.1% (R^2^_adj_) of variation which even increases to 66.6% when number of tests is *not* considered compared to decrease by 4.7% when GDP is eliminated (R^2^_adj_ = 61.4%). From this we conclude that although being strongly correlated, both GDP per capita and number of performed tests contribute to the reported incidence, however the reported COVID-19 mortality across countries is not significantly influenced by the number of tests performed *per se*.

All in all, the determined country-specific covariates which are correlated with COVID-19 mortality (GDP, press freedom, age structure of the population, time since the outbreak start, hemisphere) as well as COVID-19 incidence explain as much as about 85% of variance of mortality (R^2^ _adj_= 0.852) for the countries with sufficient number of cases and epidemy start after the 15th of February. On fig. 3 we rank the countries by mortality, original (a) as well as corrected for the abovementioned factors and incidence (b). The correction significantly alters order of the countries.

**Fig. 3.**
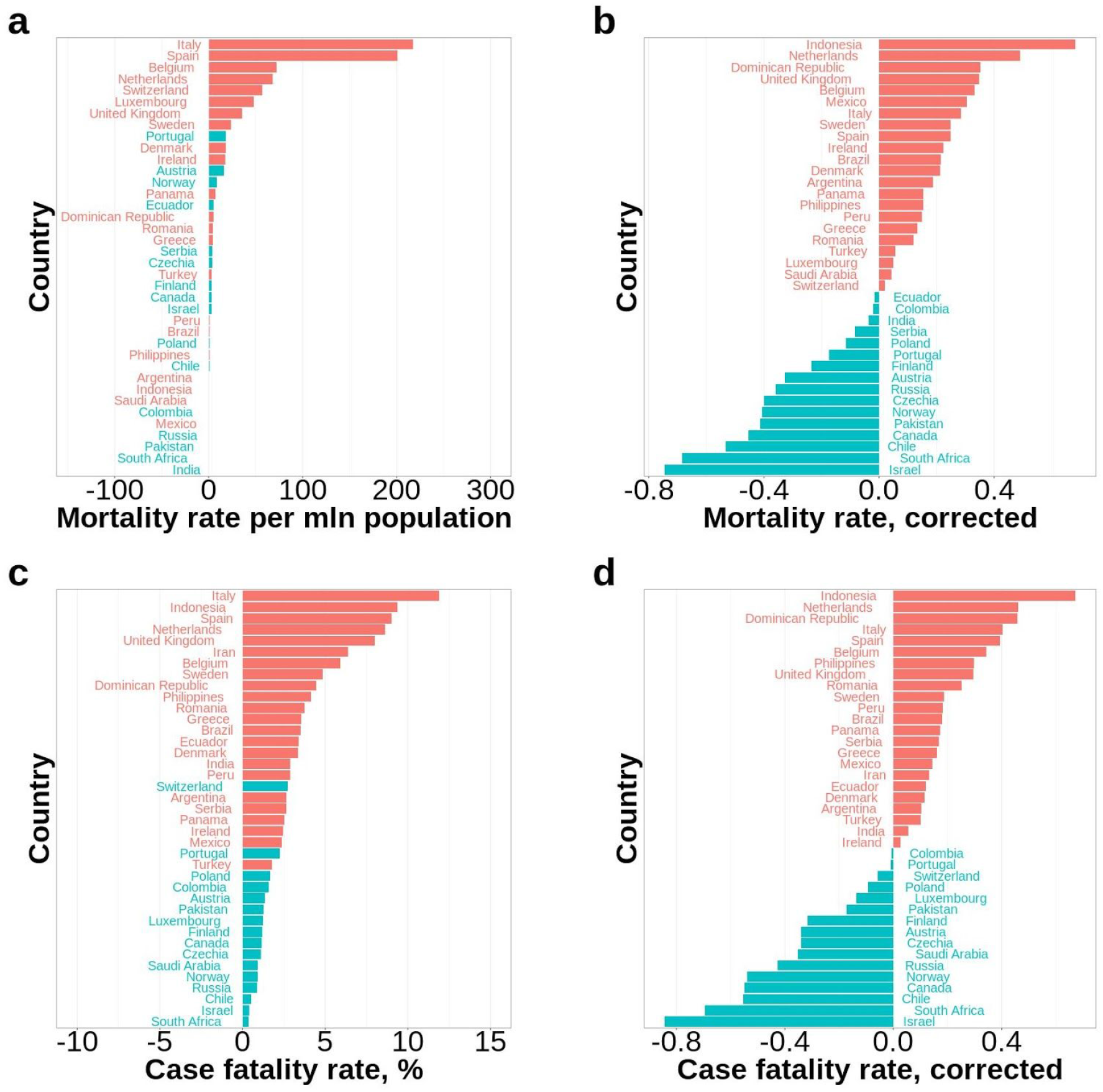
COVID-19 mortality and case fatality rate across countries as of April 1st, 2020 (outbreak start after 15th of February; at least 1000 registered cases by the 1st of April), N=39; **a**, deaths per million of people; color of the bars corresponds to relative position of the country in (b); **b**, log-transformed mortality rates corrected for COVID-19 incidence, GDP, press freedom, fraction of senior citizens, hemisphere and country-specific duration of the outbreak; **c**, case fatality rate (measured as a ratio of total deaths to total number of cases, as of April 1st, 2020), color of the bars corresponds to relative position of the country in (d); **d**, log-transformed CFRs corrected for country-specific duration of the outbreak.

### Correlation of case fatality rate with country-specific covariates

For the reason of reduction of noise that is most significant at initial stages of the outbreak, CFR has been evaluated only for the countries with at least 1000 registered COVID-19 cases during first 3 months of the year 2020. Similarly to the analysis of incidence and mortality, we worked with countries characterized by outbreak duration of no more than 40 days (N = 39 for the countries fulfilling the two requirements, see fig. 1b). For 32 out of them, number of performed tests was available. We checked correlation of all of the explanatory variables (see table 1, column 4) with case fatality rate using univariate linear regressions. Only outbreak duration demonstrated significant correlation with CFR (p = 0.028).

For further analysis, the log-transformed case fatality rate was corrected for the outbreak duration using linear model. The case fatality rates across countries are summarized on fig. 3c (original) and 3d (corrected). The total case fatality varies hugely from 0.4-0.9% in top 5 countries with lowest CFR to 8.0-11.9% in top 5 countries with the highest. We were expecting that ranking of the countries by the corrected CFR as well as ranking by mortality corrected for incidence and all other country-specific variables correlated with it should give similar results characterizing the corrected case fatality rate across countries. Indeed, scores of countries in these two rankings correlate well with each other (pearson r = 0.89, p = 1.3 × 10^−13^).

Fig. 4 illustrates change of case fatality rate during the course of pandemic for countries with highest and lowes corrected CFRs. The plot shows that the reported log-transformed CFRs gradually grow as the outbreak progresses in each country.

**Fig. 4.**
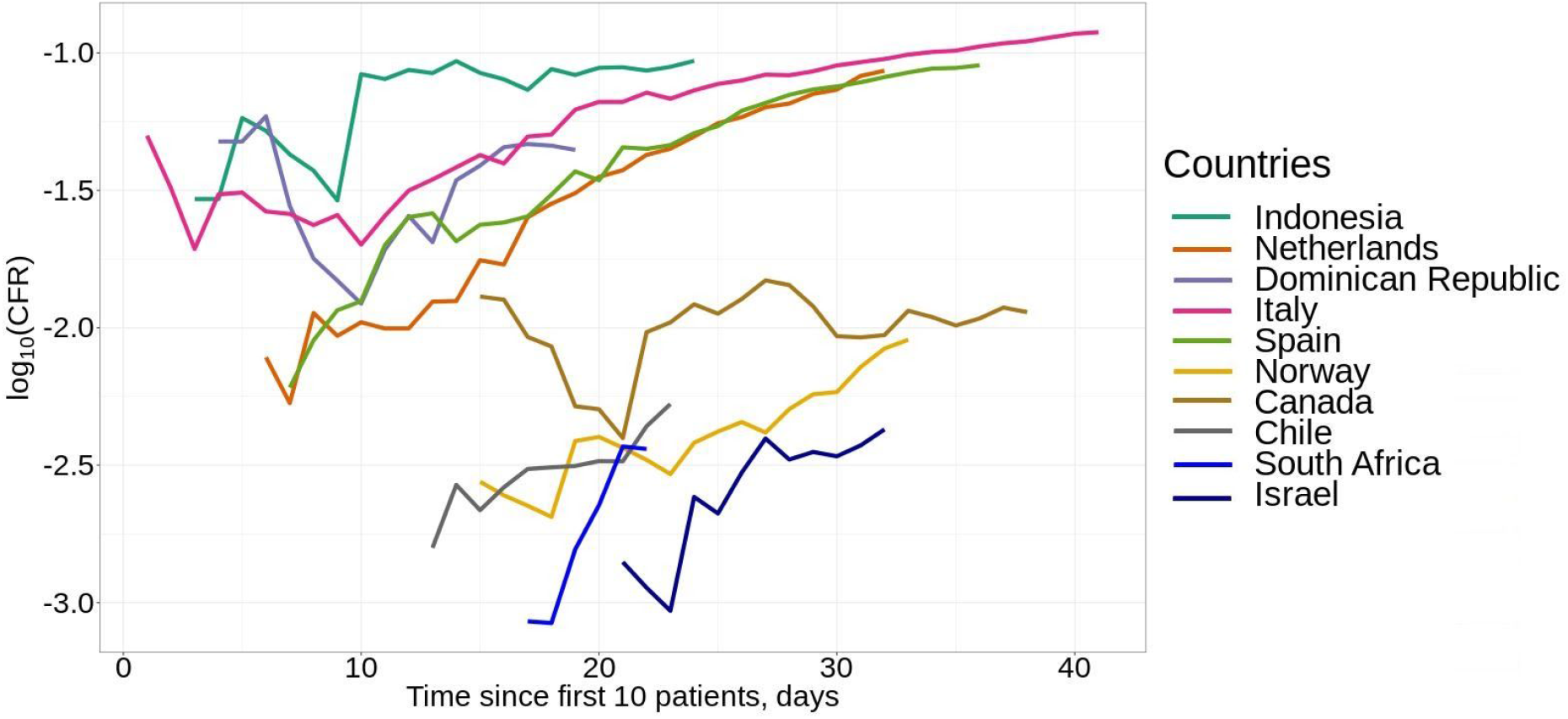
COVID-19 daily cumulative case fatality rate depending on country for top 5 countries with lowest and highest corrected values (outbreak start after 15th of February; at least 1000 registered cases by the 1st of April). The abscissa coordinate shows country-specific outbreak duration.

## Discussion

Parameters of the COVID-19 pandemic analyzed herein are important for managing and modelling the ongoing outbreak. Exploration of them during the course of the outbreak faces certain limitations based on data availability as well as standardization problems, however is crucial to timely understand spread and effect of the virus. Being a likely issue for current analysis, the anticipated data bias has also been addressed here as a phenomenon that we are aiming to untangle.

Here we show a number of correlations for the epidemiological parameters with variables characterizing economical, demographic, healthcare and political organization of the 156 countries (or overseas territories) analyzed.

Incidence, mortality and case fatality rates are expectedly correlated with duration of the COVID-19 outbreak. Other correlations demonstrated here are for incidence and mortality of the infection (CFR was herein analyzed only for 39 countries that fulfil the requirements on number of people affected and outbreak start date which substantially decreases sensitivity compared to incidence and mortality where sample sizes are much bigger).

Correlation with fraction of citizens over 65 years old is concordant with the previous findings of case fatality rate dependence on patient age (Onder, Rezza, and Brusaferro 2020). The spotted correlation of GDP per capita with incidence and mortality requires further investigation on its origins. One of the factors contributing could be higher mobility of population in ‘rich’ countries which led to quicker spread of the virus. Another one influencing the reported COVID-19 incidence (but not significantly per se influencing mortality) is the testing activity.

The other interesting finding herein is the correlation of incidence as well as mortality with freedom of the press. This supports our hypothesis that there is likely to be a practice of manipulating numbers within medical authorities of particular countries characterized by restricted press freedom and thus the numbers should be handled with caution.

We also examined possible association of incidence and mortality with country centroid latitude as well as with hemisphere it is located in. Although the correlations are evident when the linear model is done with only one explanatory variable, multiple linear regression does not support the idea of influence of latitude (p = 0.61) or hemisphere (p = 0.17) on incidence of the disease. In contrast, our analysis suggests that hemisphere might influence mortality (the latter seems to be higher in the Northern Hemisphere, p = 0.036) and thus does not not contradict the idea that outdoor temperature could impact chances for patient recovery. Finally, no significant association of any of the parameters analyzed with population density has been spotted.

Due to drastic variability in the reported COVID-19 case fatality rates across countries (from 11.9% in Italy to 0.36% in South Africa as of January-March, 2020) the practice of ranking states by CFR is tempting, but should be interpreted with extreme caution due to different testing capacities, cause-of-death assignment practices and undiagnosed COVID-19 deaths rates. Here we demonstrate that it also depends on and suggest that it should be corrected for the country-specific outbreak duration. Another approach to comparing efficiencies of healthcare systems in the times of the epidemy that we implement here is comparison of the country-specific mortalities corrected for variables that are not directly linked to healthcare service quality but have been shown to correlate with mortality: population age, wealth, press freedom, hemisphere, outbreak duration as well as for COVID-19 incidence. The two approaches give very similar results.

Abovementioned findings shed light on the course of COVID-19 pandemic and might contribute to facilitating its management across the globe.

## Data Availability

All data used for this research is publicly available

## Acknowledgements

I would like to thank Prof. Michel Georges and Prof. Yurii Aulchenko for useful discussions, ideas and extensive help with manuscript revision.

